# Implementation of a “people-like-me” tool for personalized rehabilitation after total knee arthroplasty: A mixed methods pilot study

**DOI:** 10.1101/2024.06.20.24309245

**Authors:** Jeremy Graber, Lauren Hinrichs-Kinney, Laura Churchill, Daniel D Matlock, Andrew Kittelson, Adam Lutz, Michael Bade, Jennifer Stevens-Lapsley

**Affiliations:** Eastern Colorado VA Health Care System, Geriatric Research Education and Clinical Center (GRECC), Aurora, Colorado, USA; Physical Therapy Program, Department of Physical Medicine and Rehabilitation, University of Colorado, Aurora, Colorado, USA; Active Aging Research Team, The University of British Columbia, Vancouver, British Columbia, CA; Department of Family Practice, Faculty of Medicine, The University of British Columbia, Vancouver, British Columbia, CA; Adult and Child Consortium for Health Outcomes Research and Delivery Science, University of Colorado Anschutz Medical Campus, Aurora, Colorado, USA; Division of Geriatric Medicine, Department of Medicine, University of Colorado School of Medicine, Aurora, Colorado, USA; School of Physical Therapy and Rehabilitation Science, University of Montana, Missoula, Montana, USA; ATI Physical Therapy, Greenville, South Carolina, USA; Institute for Musculoskeletal Advancement, Bolingbrook, Illinois, USA

**Keywords:** clinical decision support, total knee arthroplasty, implementation science, mixed methods, precision medicine

## Abstract

**Rationale:** While there are numerous tools available to inform if and when to use total knee arthroplasty (TKA), very few tools exist to help guide the recovery period after surgery.

**Aims and Objectives:** We piloted a decision support tool that promotes a “people-like-me” (PLM) approach to rehabilitation after total knee arthroplasty (TKA). The PLM approach encourages person-centered care by “using historical outcomes data from similar (past) patients as a template of what to expect for a new patient”. In this study, we evaluated how successfully the PLM tool was implemented and examined contextual factors that may have influenced its implementation.

**Methods:** Two outpatient physical therapy clinics (Clinics A and B) piloted the PLM tool from September 2020 – December 2022. We gathered data related to its implementation from multiple sources including the electronic health record, the tool itself, and surveys and interviews with patients and clinicians. We used an explanatory sequential mixed methods design to analyze the data overall and separately by each clinic.

**Results:** Overall, the clinics met most pre-specified implementation targets, but did not use the tool as frequently as intended. Both clinics identified time, technology, and scheduling barriers to using the tool, but Clinic A scored higher in nearly every implementation outcome. Clinic A’s success may have been related to its clinicians’ higher level of experience, more positive attitudes towards the tool, and more active approach to implementation compared to Clinic B.

**Conclusions:** The clinics met most of our implementation targets, but Clinic A experienced more success than Clinic B. Future efforts to implement this PLM tool should (1) engage clinicians as active participants in the implementation process, (2) explore whether incorporating treatment recommendations into the PLM tool and/or using alternative training strategies can enhance its ability to alter clinician behavior, (3) integrate the tool within the EHR to complement existing workflows and mitigate implementation barriers, and (4) include randomized controlled trials that evaluate the tool’s effectiveness and scalability across diverse clinical settings.

## INTRODUCTION

Total knee arthroplasty (TKA) improves quality of life for most patients with end-stage knee osteoarthritis,^1,2^ but not all patients respond predictably to TKA. About 20% of patients experience poor long-term outcomes,^3,4^ and the recovery process can be quite variable—even among those with satisfactory outcomes.^5–7^ For example, some patients quickly regain function and manage their recovery mostly independently. Other patients struggle to recover from severe impairments after surgery (e.g., pain, reduced knee mobility, weakness) and may benefit from intensive rehabilitation. Patients are also expected to recover at different rates and to different extents after surgery based on their individual characteristics.^6,7^ The variability in patients’ response to TKA suggests that postoperative care should be tailored to patient’s unique needs and goals.^6,8,9^ Yet, postoperative rehabilitation strategies are typically generic and guided by population-level evidence. In other words, most patients receive similar treatment after TKA, despite considerable variability in their preoperative prognoses and postoperative care needs.

While numerous tools have been developed to support decision making for undergoing TKA,^10,11^ no such tools exist to promote personalized rehabilitation after surgery to the best of our knowledge.^11,12^ Therefore, we developed a decision support tool to help guide decision making in rehabilitation after TKA. The tool enables a “people-like-me” (PLM) approach to rehabilitation. The PLM approach is a framework that promotes person-centered care by “using historical outcomes data from similar (past) patients as a template of what to expect for a new patient”.^6,13^ Essentially, the PLM tool generates individualized projections of TKA recovery using the historical recovery data from similar patients. We envisioned patients and clinicians could use this tool to (1) project the patient’s likely course of recovery, (2) monitor the patients’ recovery compared to similar historical patients, and (3) apply this information towards personalized care decisions.

We piloted this PLM tool in two outpatient physical therapy clinics to examine its impact on TKA rehabilitation. In this study, we evaluated the tool’s implementation using a mixed methods design informed by the Practical, Robust, Implementation and Sustainability Model (PRISM).^14^ Our goal was to understand (1) how successfully the tool was implemented and (2) the processes and contextual factors that influenced implementation at each clinic.^15^ We will use the results from this pilot study to update the tool’s features and revise our implementation strategy before deploying it on a larger scale.

## METHODS

### Description of clinical setting

The PLM tool was developed through a long-standing, collaborative relationship between researchers at the University of Colorado and ATI Physical Therapy. This relationship began in 2013 with the development of a quality improvement database of clinical TKA recovery measures. Later, members of the research and clinical teams jointly identified the need for a tool to support personalized decision making in post-TKA rehabilitation. This led to the conception of the PLM approach and, eventually, the PLM tool described in this study.

Two ATI outpatient physical therapy clinics (Clinic A and Clinic B) in the Greenville, SC, USA area piloted the PLM tool from September 2020 – December 2022. These clinics were selected based on their (1) high volume of TKA cases, (2) active contributions to the clinical database, and (3) key involvement in the PLM tool’s development, ensuring their familiarity and alignment with the tool’s objectives. All permanent, full-time physical therapy clinicians (physical therapists and physical therapist assistants) from both clinics were invited to use the tool.

### PLM tool description

The PLM tool consisted of a web-based interface that allowed clinicians to predict and monitor patient recovery using a “people-like-me” approach. Essentially, the tool used an algorithm to identify a subset of patients from a large historical database who were similar to a new patient. Then, it used the actual recovery data from these similar patients to predict the new patient’s recovery.^16,17^ These predictions were presented to patients and clinicians as “people-like-me” reference charts, which are conceptually similar to childhood growth charts. These reference charts (1) displayed the patient’s projected recovery (including the uncertainty around this projection) and (2) compared the patient’s observed recovery against this projection in terms of percentiles. To make these reference charts more interpretable, the PLM tool also provided text-based interpretations of the patient’s recovery (Figure 1). The tool generated reference charts for commonly collected outcomes after TKA including knee flexion and extension range of motion (ROM), Timed Up and Go (TUG),^18^ and the Western Ontario & McMaster Universities Arthritis Index (WOMAC) pain subscale.^19^ The tool did not provide clinicians with specific treatment recommendations. Instead, clinicians were encouraged to use their clinical judgment to determine how the information should be applied. Further information regarding the strengths and limitations of the “people-like-me” approach have been described elsewhere.^6^ We created a pared-down, open access version of the PLM tool for knee flexion and TUG recovery that can be viewed at https://cu-restore.shinyapps.io/knee_recovery_v1/.

**Figure 1.**
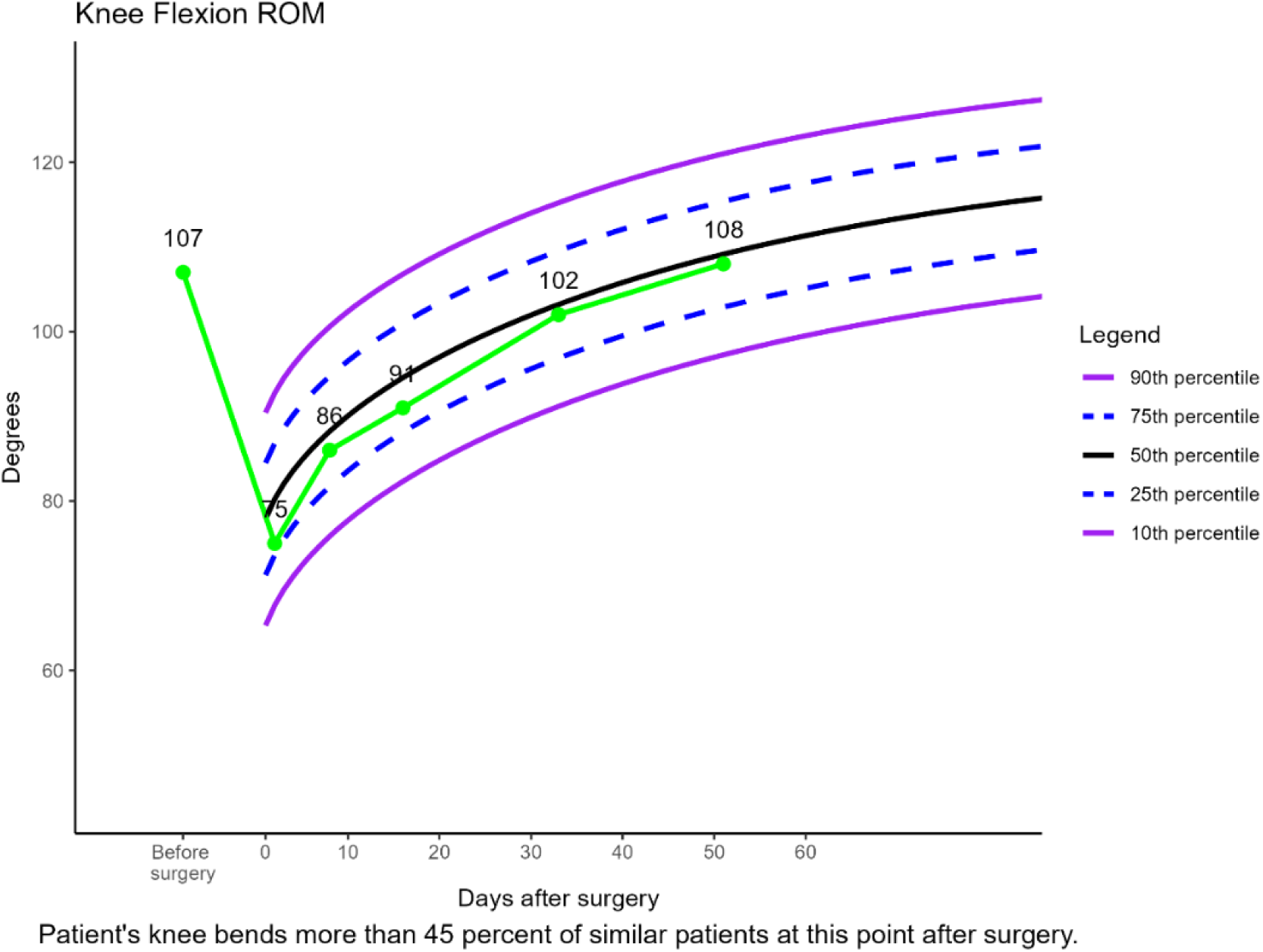
Example of “people-like-me” reference chart created by tool

### PLM tool implementation strategy

Leaders and staff members from both clinics helped design the PLM tool’s interface and strategy for implementation. Prior to launching the tool, the research team visited the clinics to describe the project’s objectives and strategy for implementation as described below. Subsequently, the research team visited the clinics yearly to re-introduce the goals of the project, troubleshoot implementation problems, and cultivate relationships with staff. The research team also met monthly with clinic leadership by videoconference to troubleshoot implementation problems and monitor the project’s progress.

Clinicians completed three self-paced, online training modules (approximately 20 minutes each) before gaining access to the tool. The training provided guidance on (1) the tool’s purpose and how to use its web-based interface, (2) how to interpret “people-like-me” reference charts and how to engage patients with them, and (3) how to use information from the tool to inform personalized decision making. Clinicians could begin using the tool immediately after completing the training. However, the clinicians completed the training at varying dates because it was self-paced, and some clinicians were hired after the pilot began.

Clinicians were provided with a fidelity checklist as part of the training. The checklist described the core components for using the tool in a patient interaction, which included collecting and entering relevant patient data, generating and printing “people-like-me” reference charts, and discussing the reference charts with the patient (Supplementary Table 1). Clinicians were strongly encouraged to adhere to the fidelity checklist and to use the tool once every 3 weeks throughout each patient’s episode of care. We selected this frequency because it mirrored the clinics’ pre-existing workflows, where clinicians were expected to assess outcomes for patients with joint replacement every 3 weeks.

Each clinic was responsible for executing the core steps described above, but they developed their own strategies for integrating them into their workflow. One local research assistant split time at each clinic supporting the tool’s implementation. The research assistant facilitated communication between the research and clinical teams, helped manage the online training, placed reminders on clinicians’ schedules to use the tool, and periodically conducted fidelity assessments during patient-clinician interactions with the tool. Additionally, the research assistant shared data (see Table 1) with the research team on a quarterly basis. The research team analyzed the data and provided each clinic with a summary of their implementation performance (i.e., audit and feedback).^20^ We provide a comprehensive summary of our implementation strategy in Supplementary Table 2 using standardized definitions from the Expert Recommendations for Implementing Change taxonomy.^20^

**Table 1.**
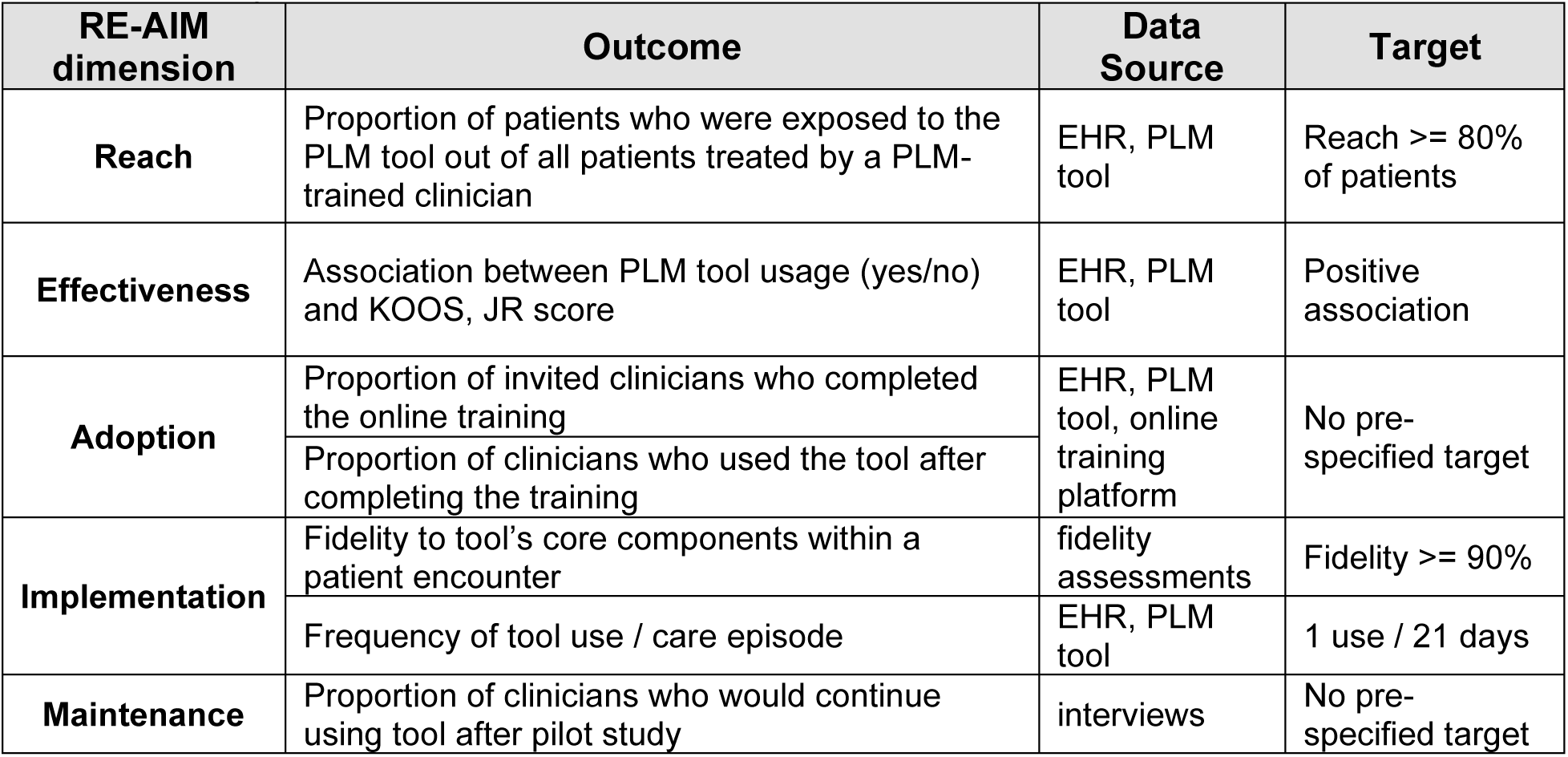
Description of RE-AIM outcomes and data sources.

**Table 2.** Description of specific PRISM contextual factors.

| PRISM Domain | Contextual Factor | Data Source |
| --- | --- | --- |
| **Organizational Characteristics** | Summary of clinicians' professional experience and training | Administrative data |
| **Organizational Perspective of Tool** | Clinicians' perceptions of tool's effectiveness, acceptability, feasibility, usability, and overall characteristics | Interviews, surveys |
| **Patient Characteristics** | Baseline characteristics of patients who were vs were not exposed to the tool out of all patients treated by a PLM-trained clinician | EHR, PLM tool |
| **Patient Perspective of Tool** | Patients' perceptions of tool's acceptability | Surveys |
|  | Patients' experiences and perceptions of using the tool during their rehabilitation | Interviews |
| **Implementation and Sustainability Infrastructure** | Implementation processes, barriers, and facilitators | Interviews, field notes |

Abbreviations: EHR = electronic health record; PLM = people-like-me.

### Data collection and outcomes

We used PRISM, a recommended framework for decision support implementation,^15^ to inform our data collection strategy. PRISM includes implementation outcomes under the dimensions of Reach, Effectiveness, Adoption, Implementation, and Maintenance (RE-AIM).^21^ We collected quantitative data corresponding to each RE-AIM dimension and pre-specified target thresholds for a subset of these outcomes (Table 1). We also collected quantitative and qualitative data related to contextual factors that may have influenced implementation. These contextual factors corresponded to the PRISM domains of Organizational Characteristics, Organizational Perspective, Patient Characteristics, Patient Perspectives, and Implementation and Sustainability Infrastructure.

We collected data from a variety of sources. Most RE-AIM outcomes were collected from the EHR, the PLM tool itself, and/or the online training platform (Table 1). We also included clinician fidelity data collected by the research assistant during onsite fidelity assessments. To examine the contextual factors that may have influenced implementation, we primarily used survey and interview data collected from patients (n=16) and clinicians (n=10) who used the tool during the pilot period. These data were also used in a companion study related to users’ perspectives of the tool,^22^ but the current analysis focused on implementation. The surveys measured patients’ and clinicians’ perceptions of the tool’s acceptability (Acceptability of Intervention Measure, AIM^23^) along with clinicians’ perceptions of the tool’s feasibility (Feasibility of Intervention Measure, FIM^23^), usability (System Usability Scale, SUS^24,25^), and other characteristics that may influence implementation (Perceived Characteristics of Intervention Scale^26^). To provide further context to the survey and interview data when needed, we also used field notes recorded by the local research assistant.

### Analysis

We used an explanatory sequential mixed methods design with a case study approach.^27^ First, we analyzed all quantitative outcomes overall and by clinic. To examine Effectiveness, we compared KOOS, JR recovery between patients treated with the PLM tool (n=167) vs patients treated before the tool became available (n=508). We used linear mixed models and adjusted for age, sex, body mass index, comorbidity profile, date of surgery, time from surgery to physical therapy evaluation, and length of outpatient physical therapy care episode. For all other quantitative outcomes, we calculated descriptive statistics and tested for between-group differences when indicated (e.g., comparing patients who were vs were not reached). Due to the quantitative differences we found between clinics, we decided to integrate our data sources using a case study approach. We used the qualitative interview data to help explain and provide context to the quantitative similarities and differences observed between clinics.

Two members of the research team (LC and JG) analyzed the clinician interview data using descriptive content analysis with a blend of deductive and inductive approaches.^28,29^ First, they developed a preliminary codebook based on PRISM domains. Next, they jointly coded the first 4 interviews and subsequently revised the codebook by incorporating inductive codes. Both coders independently re-coded the first 4 interviews and then reviewed them together to ensure consistency. The final 6 interviews were coded independently by 1 of the 2 coders. Throughout this process, the coders consulted with a PhD-trained qualitative researcher for methodological guidance and to resolve any coding disagreements. All coding procedures were conducted using Dedoose software (version 9.0.90). Once all interviews had been coded, we entered the data into a matrix and re-examined the codes by clinic location.^30^ This allowed us to detect similarities and differences between clinics. Finally, we used a narrative format to report our findings by RE-AIM dimension. We organized contextual data by PRISM domain to help explain and enhance our understanding of the RE-AIM outcomes.^27^

## RESULTS

The quantitative RE-AIM outcomes are displayed in Table 3. Collectively, the clinics met or exceeded targets related to Reach, Effectiveness, and fidelity, but they did not use the tool as frequently as recommended. Clinic A scored higher in nearly every outcome and met more targets than Clinic B.

**Table 3.**
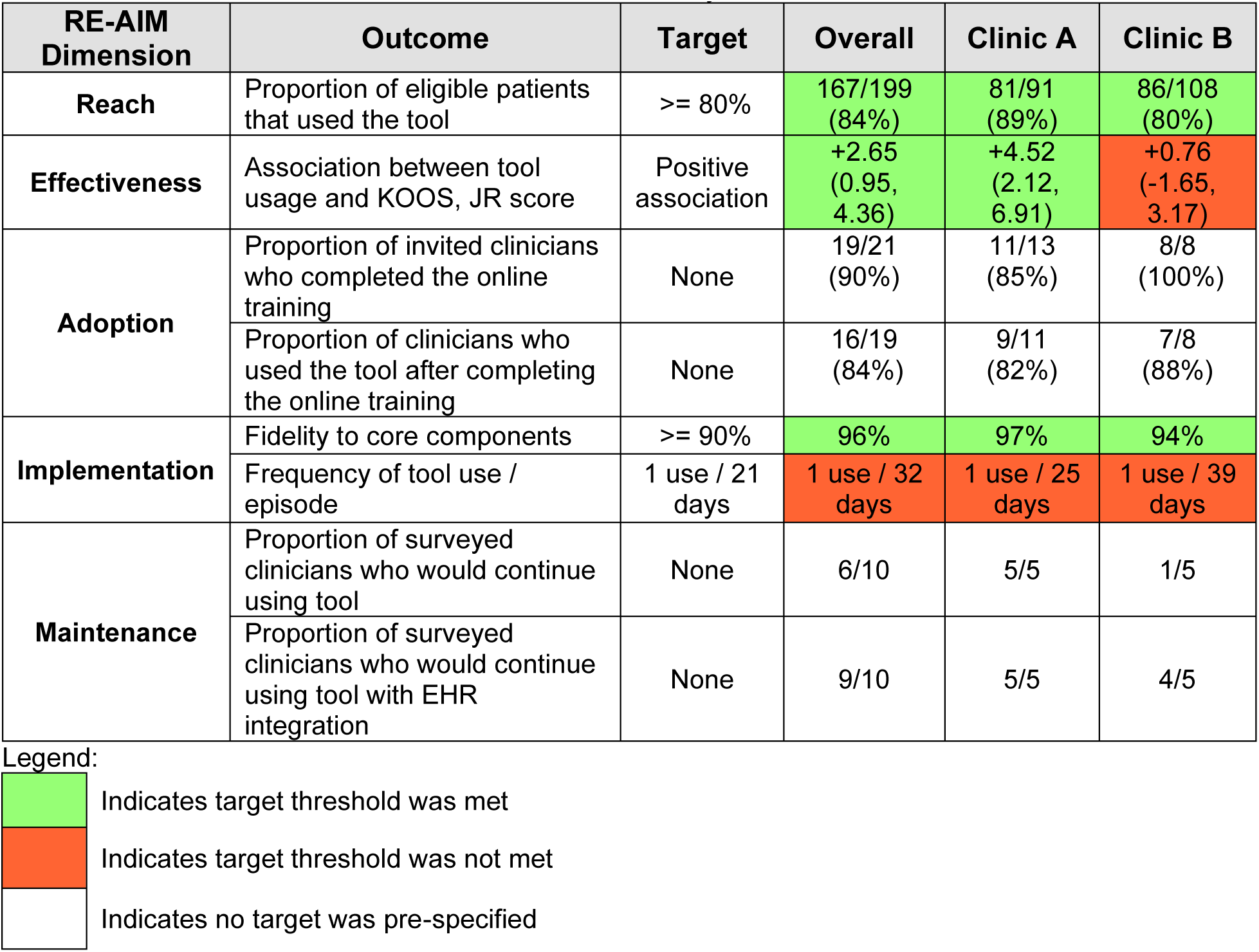
RE-AIM outcomes summarized overall and by clinic.

| RE-AIM Dimension | Outcome | Target | Overall | Clinic A | Clinic B |
| --- | --- | --- | --- | --- | --- |
| Reach | Proportion of eligible patients that used the tool | $\geq 80\%$ | 167/199<br>(84%) | 81/91<br>(89%) | 86/108<br>(80%) |
| Effectiveness | Association between tool usage and KOOS, JR score | Positive association | +2.65<br>(0.95, 4.36) | +4.52<br>(2.12, 6.91) | +0.76<br>(-1.65, 3.17) |
| Adoption | Proportion of invited clinicians who completed the online training | None | 19/21<br>(90%) | 11/13<br>(85%) | 8/8<br>(100%) |
|  | Proportion of clinicians who used the tool after completing the online training | None | 16/19<br>(84%) | 9/11<br>(82%) | 7/8<br>(88%) |
| Implementation | Fidelity to core components | $\geq 90\%$ | 96% | 97% | 94% |
|  | Frequency of tool use / episode | 1 use / 21 days | 1 use / 32 days | 1 use / 25 days | 1 use / 39 days |
| Maintenance | Proportion of surveyed clinicians who would continue using tool | None | 6/10 | 5/5 | 1/5 |
|  | Proportion of surveyed clinicians who would continue using tool with EHR integration | None | 9/10 | 5/5 | 4/5 |

Legend:
|  |  |
| --- | --- |
|  | Indicates target threshold was met |
|  | Indicates target threshold was not met |
|  | Indicates no target was pre-specified |

The PRISM contextual data are displayed in Table 4. Clinic A was larger, and its clinicians had more experience, longer tenure in their organization, and more advanced training than Clinic B. Clinic A clinicians also had more favorable perspectives of the tool than Clinic B, but patient perspectives at both clinics were mostly positive. Clinicians identified common barriers and facilitators to using the tool, but the clinics described different processes for integrating it within their workflows.

**Table 4.** PRISM contextual factors summarized overall and by clinic.

| **PRISM Domain** | **Outcome** | **Overall** | **Clinic A** | **Clinic B** |
| --- | --- | --- | --- | --- |
| **Organizational Characteristics** | Full-time clinicians (FTE equivalent) | 25 | 17 | 8 |
|  | Residency-trained clinicians | 5 | 5 | 0 |
|  | Board-certified clinicians (e.g., OCS) | 3 | 2 | 1 |
|  | Clinicians' average years of experience | 4 | 6 | 2 |
|  | Clinicians' average years of experience in organization | 4 | 6 | 2 |
| **Organizational Perspective of Tool** | Acceptability of Intervention Measure (AIM) | 3.7 / 5 | 4.1 / 5 | 3.4 / 5 |
|  | Feasibility of Intervention Measure (FIM) | 3.9 / 5 | 4.1 / 5 | 3.8 / 5 |
|  | System Usability Scale (SUS) | 66.5 / 100 | 71.5 / 100 | 61.5 / 100 |
|  | Perceived Characteristics of Intervention Scale (PCIS) | 3.7 / 5 | 3.8 / 5 | 3.6 / 5 |
|  | Qualitative perception of tool's value | Useful for patient education | Useful for most patients | Useful for a subset of patients |
| **Patient Characteristics** | Characteristics of reached vs unreached patients | See Table 5 |  |  |
| **Patient Perspective of tool** | Acceptability of Intervention Measure (AIM) | 4.4 / 5 | 4.4 / 5 | 4.3 / 5 |
|  | Qualitative perception of tool's value | Useful for increasing motivation and managing expectations |  |  |
| **Implementation and Sustainability Infrastructure** | Barriers | • Time<br>• Technology<br>• Scheduling |  | • Reliance on support staff |
|  | Facilitators | • Simple interface<br>• Data collection complements workflows | • Confidence ↑ with experience<br>• Preparation before patient arrives | • Reliance on support staff |
|  | Implementation approach | Support from research team | Active clinician involvement | Passive clinician involvement |

Abbreviations: FTE = full time equivalent, OCS = orthopedic clinical specialist

### Reach

Overall, 84% of eligible patients had the PLM tool used to inform their care. A higher proportion of patients were reached at Clinic A vs Clinic B, although the difference was not statistically significant (89% vs 80%, p = 0.07). The characteristics of the reached vs unreached patients are available in Table 5. The unreached group had a higher proportion of females (75% vs 53%, p = 0.02) and earlier surgical dates (median: 05/01/2021 vs 12/23/2021, p = 0.005) compared to the reached group. Clinicians at both clinics identified barriers (described under implementation below) that sometimes prevented them from using the tool with patients. At Clinic A, clinicians described how these barriers improved over time. For example, one clinician described how they identified and addressed a scheduling issue related to patients with Medicare, which improved the tool’s reach at their clinic moving forward.

> *After we identified why we seemed to be missing some people, and they were all Medicare, we kind of put the pieces together. After we discovered that, there didn’t seem to be any issues with using it.*

> *-Clinician 7, Clinic A*

**Table 5.**
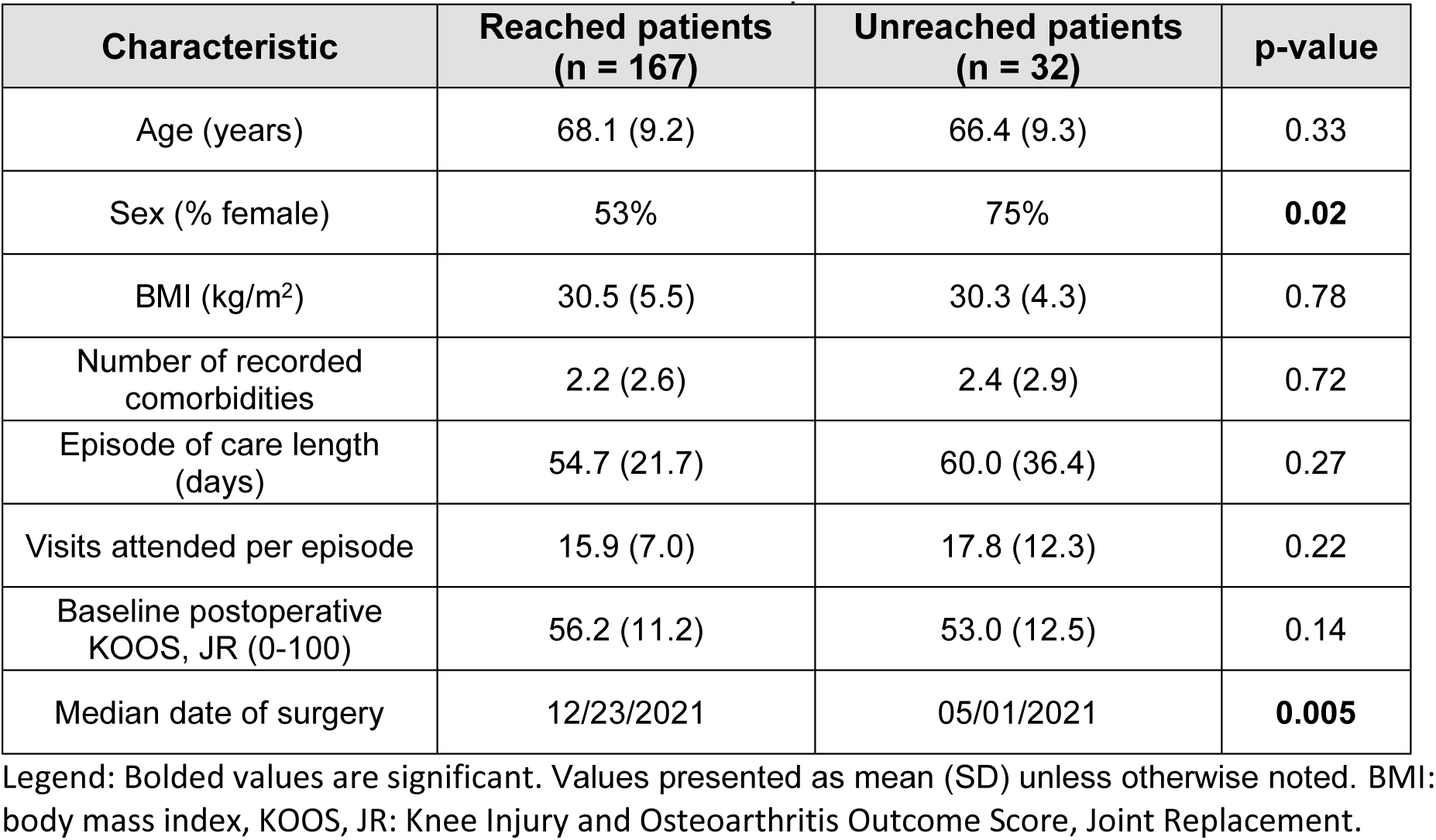
Characteristics of reached vs unreached patients.

No clinicians mentioned anything directly related to using the tool less often with female patients. However, two clinicians mentioned rare cases when they chose not to share the tool’s information with patients who were experiencing a challenging recovery, because they were concerned it could be discouraging or demotivating.^22^

### Effectiveness

PLM tool usage was associated with improved patient-reported knee health (KOOS, JR), and this association was greater within Clinic A than within Clinic B (+4.52 points vs +0.76 points, p = 0.03). At both clinics, patients rated the tool as acceptable, and clinicians and patients felt the tool was effective for educating, motivating, and reassuring patients during their recovery from TKA.^22^ However, most clinicians stated they did not use the tool to adjust or personalize treatments, because they felt confident in their own ability to determine patients’ prognoses and monitor their recovery.

> *I have my own parameters in my head for what my expectations are for them…*

> *-Clinician 2, Clinic B*

At Clinic B, most clinicians mentioned that some patients seemed disinterested in the tool, or that the tool was only effective for a subgroup of patients who found the tool’s information interesting. Several of these clinicians described using the tool with patients in a way that seemed unenthusiastic or perfunctory.

> *I do wonder because I feel like with the wins, like the huge like, “oh, my gosh, this is so awesome!” was a subgroup. I wonder how many of those it takes to be like to be overall successful? So like, what’s your number to treat?*

> *-Clinician 1, Clinic B*

> *I was like, “OK. I’m just gonna give you these [reference chart printouts]. If you want to look at them, you look at them, but some of them honestly just didn’t really care. Some of them were right on track and were going, “Oh, that’s nice. That’s cool.” So*.

> *-Clinician 8, Clinic B*

Clinic A clinicians similarly perceived that some patients were more interested in the tool than others. However, they generally described how the tool facilitated useful conversations with patients and provided visual reinforcement for patient education.

> *I feel like most people were pretty interested in it. Just having the sheet, I feel like was a good thing. But most people are visual kind of learners.*

> *-Clinician 7, Clinic A*

In contrast to the approach described by several Clinic B clinicians, one Clinic A clinician felt that their enthusiasm for using the tool enhanced its impact on patients.

> *So, my excitement about “this is where you are, and I can see it, and I can compare it”, probably translates to the way I educate about it. I think it was definitely useful for my personality in the way I like to interact with my patients, and that truthfully plays a big role in what they take away. So, if I don’t care and I’m telling them this data, they’re not gonna care.*

> *-Clinician 6, Clinic A*

### Adoption

We observed high rates of adoption at both clinics as all permanent, full-time clinicians were strongly encouraged by leadership to use the tool. The only clinicians who did not complete the training or use the tool moved clinic locations during the pilot study period. The median time to complete the self-paced training was 37 days after initial assignment, with no significant between-clinic difference.

### Implementation

#### Common Barriers and Facilitators

Both clinics demonstrated high fidelity within individual patient encounters, but the tool was used less frequently than intended. Clinicians reported limited time was the biggest barrier to using the tool consistently at both clinics.

> *It’s true, in the whole scheme of things is three minutes too much? No, but when you’re already struggling to maintain your workload, just the thought of an extra three minutes was tough.*

> *-Clinician 1, Clinic B*

Many clinicians reported that time limitations were sometimes worsened by technological issues. For example, sometimes the tool loaded slowly and other times the printers were not working. Clinicians also stated that scheduling issues made it challenging to remember when to use the tool, like when patients alternated between clinicians. Many clinicians reported the tool’s interface was simple and easy to use. Several also reported the outcome measures required to use the tool complemented their clinics’ existing workflows and practice patterns.

#### Differences in Implementation

Clinic A used the tool more frequently per patient (1 use / 25 days vs 1 use / 39 days, p < 0.001) and performed more fidelity assessments (30 vs 5) than Clinic B. Clinic A clinicians also perceived the tool more positively; they felt it was more feasible (4.1 vs 3.8, FIM) and usable (73.1 vs 61.5, SUS) than Clinic B. These findings may be partially explained by the clinics’ contrasting approaches to implementation.

At Clinic B, clinicians described relying heavily on their support staff for implementation. Clinicians discussed the reference charts with patients, but their support staff were responsible for all other tasks (i.e., using the tool’s web-based interface, printing the reference charts, keeping track of when to use tool). Clinicians described this as both a facilitator and a barrier. For example, they mentioned it reduced the time required to use the tool.

> *I made it clear to my supervisors that I wasn’t interested in taking on more responsibility with respect to that. I was like, I’ll collect the data, I’ll read them the graphs, but I do not want to use the [tool]…I just don’t have time for that, and we actually have support staff that do….*

> *-Clinician 2, Clinic B*

However, several Clinic B clinicians stated they never became comfortable using the tool because of their reliance on support staff. Other clinicians made comments that suggested they did not completely understand how the tool functioned.

> *I found it a little bit hard to figure out how to get in there and get to the right patient, get the info in, but honestly, I didn’t have a whole lot of practice with that because we did start passing those off pretty quick to different staff members.*

> *-Clinician 9, Clinic B*

Clinic B clinicians also stated their support staff were very busy, which sometimes meant they were unavailable to help use the tool if unanticipated issues arose. The research assistant also noted the tool was not used at consistent intervals at Clinic B, which made it challenging to conduct fidelity assessments.

> *And so even with our [support staff] staff taking the reins…it still felt a little bit stressful. And sometimes, just because we weren’t sure if the graphs were, if the data was printing out right or if the right points were on there. And sometimes we would have issues printing and so it was a little bit of a stress. So, I stopped telling patients we were gonna give them those because I was like, “oh, I don’t know if we’re actually going to get them.”*

> *-Clinician 9, Clinic B*

Clinic A clinicians described taking a more active role in the tool’s implementation, where they used the tool independently with occasional help from support staff. Most clinicians explained they gained confidence using the tool over time.

> *I think it was one of those things, like, you kind of gotta use…once you do a couple of live cases it was pretty easy*

> *-Clinician Interview 5, Clinic A*

Clinic A clinicians frequently explained the tool became part of their standard care for patients with TKA. Several clinicians mentioned they prepared to use the tool before a patient arrived, which helped them overcome any barriers that arose. Other clinicians said using the tool became so routine that they could integrate it seamlessly within a treatment session.

> *It just was a component of what we did. Maybe I sprinkled it into rest breaks here and there. Just like intentional timing of when I did it helped me. But, I really didn’t find it hard to incorporate into an hour to an hour and 15-minute session. There was plenty of opportunity to do so.*

> *-Clinician 6, Clinic A*

Clinicians also described making adaptations to the tool as they gained confidence. For example, some clinicians made visual changes to the printed reference charts to make them more interpretable for patients. One clinician described how they gradually adapted their strategy over time to maximize the tool’s usefulness for patients.

> *I think it was probably more cookie cutter when I first started [using the tool], and I melded it into what I actually wanted as I went…I think that was probably an evolution for me of learning how to maximize the benefit from it and not just make it empty words for a patient.*

> *-Clinician 6, Clinic A*

### Maintenance

All Clinic A clinicians stated they would choose to continue using the tool after the pilot compared to only 1/5 clinicians at Clinic B. Clinic A also rated the tool as more acceptable (4.1 vs 3.4, AIM) and more favorable overall (3.8 vs 3.6, PCIS). Clinic A clinicians stated they would continue using the tool because it helped them with patient education and provided tangible value to their patients. At Clinic B, most clinicians did not feel the tool provided enough value to keep using, given the time and energy it required.

> *I do like the idea of it, but I’ll be honest. I just don’t think it’s a feasible thing.*

> *-Clinician 9, Clinic B*

Several Clinic B clinicians felt the tool only provided incremental value beyond their usual patient education interventions, while others reiterated the tool was most effective with only certain patients (as described under Effectiveness). However, 4/5 Clinic B clinicians stated they would use the tool if it was integrated in the EHR, which they felt would make it more feasible.

## DISCUSSION

In this pilot study, we evaluated the implementation of a new PLM tool that promotes a personalized, “people-like-me” (PLM) approach to rehabilitation after TKA. We used a package of strategies (Supplementary Table 2) to support the tool’s implementation in two outpatient physical therapy clinics. The clinics met targets related to Reach, Effectiveness, and fidelity, but they did not use the tool as frequently as recommended. Clinic A scored higher in nearly every outcome and met more implementation targets than Clinic B. Clinicians’ perspective of the tool also differed by clinic, and they used different strategies for integrating it within their workflows. The rich results of this mixed methods study will help guide future efforts to implement the tool more widely.

Clinic A’s active approach to implementation seemed to account for much of their success. Clinic A clinicians described the tool as part of their usual care for patients with TKA. Therefore, they expected to use it at regular intervals, and they assumed most of the responsibility for integrating it within their workflow. They reported feeling confident using the tool, which may have empowered them to overcome barriers and adapt their strategies for using it over time. In contrast, Clinic B clinicians described taking a more passive role in implementation and seemed to be less comfortable using the tool. As a result, they appeared to be less prepared to overcome implementation barriers, which may have negatively impacted their perception of the tool’s feasibility and usability. Future efforts to implement the tool should facilitate clinicians’ active participation. These efforts could include supporting clinicians with interactive implementation assistance or providing clinicians with individual feedback on their implementation performance.^20,31^

Clinic A’s more positive perspective of the tool likely also influenced their implementation success.^32–34^ Clinicians are more likely to use interventions that are perceived to address gaps in patient care or provide advantages over usual care.^14,15,35^ Most clinicians felt the tool enhanced their patient education, but several Clinic B clinicians felt this enhancement was marginal or only applicable to a subset of patients. These clinicians’ perceptions of the tool may have been improved if they were provided with (a) evidence regarding its positive impact on patients’ outcomes and experiences^22^ and/or (b) information on how it may improve their work experience.

Most Clinic A and B clinicians stated they did not use the tool to adjust or personalize their treatment plans, because they did not perceive this to be a gap in patient care. They described feeling confident in their own ability to provide personalized treatment after TKA. Therefore, new training and implementation strategies may be needed for the tool to influence clinicians’ beliefs and behaviors. This could include developing training strategies that incorporate more active learning techniques such as peer review, performance feedback, and mentored patient interactions.^36,37^ Alternatively, the tool could be updated to more effectively prompt changes in clinician behavior. Clinicians may have been more likely to change/adjust their treatment plan if the tool provided a specific recommendation for their consideration.^38,39^ For instance, if the tool identified that a patient’s knee flexion ROM was recovering more slowly than expected, it could recommend an evidence-based exercise to improve their motion.^40^ This approach could potentially enhance the tool’s effectiveness and reduce variability in how clinicians use it.

There were differences in organizational characteristics between Clinics A and B that may have impacted our findings. At Clinic A, clinicians had more experience, longer tenure in their organization, and more advanced training on average compared to Clinic B. Therefore, clinicians may have been better equipped to implement new treatment approaches and overcome implementation barriers. Clinic A is also well known within their organization for their highly-collaborative, team-based culture, which may have provided a more supportive implementation environment.^14^ We also cannot rule out whether the clinics differed in other unmeasured characteristics (e.g., readiness for change) that influenced implementation. Regardless, our results suggest that clinics will vary in their ability to overcome implementation barriers (even within the same organization and geographical area), so commonly identified barriers should be proactively addressed.

Clinics A and B identified common infrastructural barriers to using the tool including time, scheduling, and technological issues. These issues frequently hinder decision support implementation,^33,41^ and there are many recommended strategies to overcome them.^32,42–45^ However, integrating the tool within the electronic health record (EHR) would likely be the most effective next step. EHR integration would eliminate redundant data entry and embed the tool directly within existing workflows. Nearly all participating clinicians believed EHR integration would improve their experience using the tool, and most Clinic B clinicians stated it would persuade them to continue using it in the future.

Clinicians did not specifically mention the costs associated with implementing the PLM tool in this study, but cost will be an important determinant of the tool’s future uptake.^14,15^ The primary implementation costs for this pilot study were salary support for the research assistant and the time burden associated with using the PLM tool. We anticipate different costs will be associated with future efforts to implement the tool. For example, integrating the tool within the EHR will require non-trivial startup costs, but will also reduce the burden of data entry and the need for dedicated support staff. Future studies of this tool’s implementation should examine these costs in relation to the tool’s benefits (e.g., impact on patient outcomes and satisfaction) using formal cost-benefit analyses.

### Limitations

This study does have some limitations. We only piloted the PLM tool in two clinics from the same clinic system and geographical region, and our sample size for the clinician interviews and surveys was small. Our results may not generalize to clinics with different characteristics or clinics from different health care systems or regions. We also did not capture all baseline clinic characteristics that could have influenced implementation, which could have explained some of the differences we observed between Clinics A and B. Nonetheless, our study’s mixed methods design leveraged multiple data sources and followed an established outcomes framework, which allowed for a robust examination of the tool’s implementation.

### Conclusion

We piloted a new tool that promotes a personalized, “people-like-me” (PLM) approach to post-TKA rehabilitation in two outpatient physical therapy clinics. The clinics achieved implementation targets related to Reach, Effectiveness, and fidelity, but used the tool less frequently than recommended. Notably, Clinic A scored higher in nearly every outcome, and its clinicians viewed the tool more favorably than Clinic B. Based on the results of this study, future research involving this PLM tool should (1) engage clinicians as active participants in the implementation process, (2) explore whether incorporating treatment recommendations into the tool and/or using alternative training strategies can enhance its ability to alter clinician behavior, and (3) integrate the tool within the EHR to complement existing workflows and mitigate implementation barriers, and (4) include randomized controlled trials that evaluate the tool’s effectiveness and scalability across diverse clinical settings.

## Data Availability

All data produced in the present study are available upon reasonable request to the authors

## Abbreviations

PLM: “people-like-me”
TKA: total knee arthroplasty

## REFERENCES

1. Shan L, Shan B, Suzuki A, Nouh F, Saxena A. Intermediate and long-term quality of life after total knee replacement: A systematic review and meta-analysis. J Bone Joint Surg Am. Jan 21 2015;97(2):156–68. doi:10.2106/JBJS.M.00372

2. Ethgen O, Bruyere O, Richy F, Dardennes C, Reginster JY. Health-related quality of life in total hip and total knee arthroplasty. A qualitative and systematic review of the literature. J Bone Joint Surg Am. May 2004;86(5):963–74. doi:10.2106/00004623-200405000-00012

3. Kahlenberg CA, Nwachukwu BU, McLawhorn AS, Cross MB, Cornell CN, Padgett DE. Patient satisfaction after total knee replacement: A systematic review. HSS J. Jul 2018;14(2):192–201. doi:10.1007/s11420-018-9614-8

4. Beswick AD, Wylde V, Gooberman-Hill R, Blom A, Dieppe P. What proportion of patients report long-term pain after total hip or knee replacement for osteoarthritis? A systematic review of prospective studies in unselected patients. BMJ Open. 2012;2(1):e000435. doi:10.1136/bmjopen-2011-000435

5. Kittelson AJ, Elings J, Colborn K, et al. Reference chart for knee flexion following total knee arthroplasty: A novel tool for monitoring postoperative recovery. BMC Musculoskelet Disord. Jul 22 2020;21(1):482. doi:10.1186/s12891-020-03493-x

6. Kittelson AJ, Hoogeboom TJ, Schenkman M, Stevens-Lapsley JE, van Meeteren NLU. Person-centered care and physical therapy: A “people-like-me” approach. Phys Ther. Jan 23 2020;100(1):99–106. doi:10.1093/ptj/pzz139

7. Wylde V, Penfold C, Rose A, Blom AW. Variability in long-term pain and function trajectories after total knee replacement: A cohort study. Orthop Traumatol Surg Res. Nov 2019;105(7):1345–1350. doi:10.1016/j.otsr.2019.08.014

8. Weiss JM, Noble PC, Conditt MA, et al. What functional activities are important to patients with knee replacements? Clinical Orthopaedics and Related Research. 2002;404:172–188.

9. Tilbury C, Haanstra TM, Leichtenberg CS, et al. Unfulfilled expectations after total hip and knee arthroplasty surgery: There is a need for better preoperative patient information and education. J Arthroplasty. Oct 2016;31(10):2139–45. doi:10.1016/j.arth.2016.02.061

10. Pacheco-Brousseau L, Stacey D, Desmeules F, et al. Instruments to assess appropriateness of hip and knee arthroplasty: A systematic review. Osteoarthritis Cartilage. Jul 2023;31(7):847–864. doi:10.1016/j.joca.2023.02.077

11. van der Sluis G, Jager J, Punt I, et al. Current status and future prospects for shared decision making before and after total knee replacement surgery-a scoping review. Int J Environ Res Public Health. Jan 14 2021;18(2)doi:10.3390/ijerph18020668

12. Cochrane JA, Flynn T, Wills A, Walker FR, Nilsson M, Johnson SJ. Clinical decision support tools for predicting outcomes in patients undergoing total knee arthroplasty: A systematic review. J Arthroplasty. May 2021;36(5):1832–1845 e1. doi:10.1016/j.arth.2020.10.053

13. Alemi F, Erdman H, Griva I, Evans CH. Improved statistical methods are needed to advance personalized medicine. Open Transl Med J. Jan 1 2009;1:16–20. doi:10.2174/1876399500901010016

14. Feldstein AC, Glasgow RE. A practical, robust implementation and sustainability model (prism) for integrating research findings into practice. Jt Comm J Qual Patient Saf. Apr 2008;34(4):228–43. doi:10.1016/s1553-7250(08)34030-6

15. Trinkley KE, Kahn MG, Bennett TD, et al. Integrating the practical robust implementation and sustainability model with best practices in clinical decision support design: Implementation science approach. Original Paper. J Med Internet Res. Oct 29 2020;22(10):e19676. doi:10.2196/19676

16. van Buuren S. Curve matching: A data-driven technique to improve individual prediction of childhood growth. Ann Nutr Metab. 2014;65(2-3):227–33. doi:10.1159/000365398

17. Kim C, Colborn KL, van Buuren S, Loar T, Stevens-Lapsley JE, Kittelson AJ. Neighbors-based prediction of physical function after total knee arthroplasty. Sci Rep. Aug 18 2021;11(1):16719. doi:10.1038/s41598-021-94838-6

18. Podsiadlo D, Richardson S. The timed “up & go“: A test of basic functional mobility for frail elderly persons. J Am Geriatr Soc. Feb 1991;39(2):142–8. doi:10.1111/j.1532-5415.1991.tb01616.x

19. McConnell S, Kolopack P, Davis AM. The western ontario and mcmaster universities osteoarthritis index (womac): A review of its utility and measurement properties. Arthritis and rheumatism. Oct 2001;45(5):453–61. doi:10.1002/1529-0131(200110)45:5<453::aid-art365>3.0.co;2-w

20. Powell BJ, Waltz TJ, Chinman MJ, et al. A refined compilation of implementation strategies: Results from the expert recommendations for implementing change (eric) project. Implement Sci. Feb 12 2015;10(1):21. doi:10.1186/s13012-015-0209-1

21. Glasgow RE, Harden SM, Gaglio B, et al. Re-aim planning and evaluation framework: Adapting to new science and practice with a twenty-year review. Frontiers in public health. 2019;7:64.

22. Churchill L, Graber J, Mealer M, et al. Patient and clinician perceptions of a “people-like-me” tool for personalized rehabilitation after total knee arthroplasty: A qualitative interview study. medRxiv. Preprint posted online June 15, 2024 2024;doi:10.1101/2023.10.23.23297404

23. Weiner BJ, Lewis CC, Stanick C, et al. Psychometric assessment of three newly developed implementation outcome measures. Implement Sci. Aug 29 2017;12(1):108. doi:10.1186/s13012-017-0635-3

24. Brooke J. Sus-a quick and dirty usability scale. Usability evaluation in industry. 1996;189(194):4–7.

25. Lewis JR. The system usability scale: Past, present, and future. International Journal of Human–Computer Interaction. 2018;34(7):577–590.

26. Cook JM, Thompson R, Schnurr PP. Perceived characteristics of intervention scale: Development and psychometric properties. Assessment. Dec 2015;22(6):704–14. doi:10.1177/1073191114561254

27. Fetters MD, Curry LA, Creswell JW. Achieving integration in mixed methods designs-principles and practices. Health Serv Res. Dec 2013;48(6 Pt 2):2134–56. doi:10.1111/1475-6773.12117

28. DeCuir-Gunby JT, Marshall PL, McCulloch AW. Developing and using a codebook for the analysis of interview data: An example from a professional development research project. Field Method. May 2010;23(2):136–155. doi:10.1177/1525822x10388468

29. Hsieh HF, Shannon SE. Three approaches to qualitative content analysis. Qual Health Res. Nov 2005;15(9):1277–88. doi:10.1177/1049732305276687

30. Gale NK, Heath G, Cameron E, Rashid S, Redwood S. Using the framework method for the analysis of qualitative data in multi-disciplinary health research. BMC Med Res Methodol. Sep 18 2013;13(1):117. doi:10.1186/1471-2288-13-117

31. Waltz TJ, Powell BJ, Matthieu MM, et al. Use of concept mapping to characterize relationships among implementation strategies and assess their feasibility and importance: Results from the expert recommendations for implementing change (eric) study. Implement Sci. Aug 7 2015;10:109. doi:10.1186/s13012-015-0295-0

32. Van de Velde S, Kunnamo I, Roshanov P, et al. The guides checklist: Development of a tool to improve the successful use of guideline-based computerised clinical decision support. Implement Sci. Jun 25 2018;13(1):86. doi:10.1186/s13012-018-0772-3

33. Devaraj S, Sharma SK, Fausto DJ, Viernes S, Kharrazi H. Barriers and facilitators to clinical decision support systems adoption: A systematic review. Journal of Business Administration Research. 2014;3(2):36.

34. Greenes RA, Bates DW, Kawamoto K, Middleton B, Osheroff J, Shahar Y. Clinical decision support models and frameworks: Seeking to address research issues underlying implementation successes and failures. J Biomed Inform. Feb 2018;78:134–143. doi:10.1016/j.jbi.2017.12.005

35. Greenhalgh T, Robert G, Macfarlane F, Bate P, Kyriakidou O. Diffusion of innovations in service organizations: Systematic review and recommendations. Milbank Q. 2004;82(4):581–629. doi:10.1111/j.0887-378X.2004.00325.x

36. Leahy E, Chipchase L, Calo M, Blackstock FC. Which learning activities enhance physical therapist practice? Part 2: Systematic review of qualitative studies and thematic synthesis. Physical Therapy. 2020;100(9):1484–1501. doi:10.1093/ptj/pzaa108

37. Leahy E, Chipchase L, Calo M, Blackstock FC. Which learning activities enhance physical therapist practice? Part 1: Systematic review and meta-analysis of quantitative studies. Physical therapy. 2020;100(9):1469–1483.

38. Kawamoto K, Houlihan CA, Balas EA, Lobach DF. Improving clinical practice using clinical decision support systems: A systematic review of trials to identify features critical to success. BMJ. Apr 2 2005;330(7494):765. doi:10.1136/bmj.38398.500764.8F

39. Kilsdonk E, Peute LW, Jaspers MW. Factors influencing implementation success of guideline-based clinical decision support systems: A systematic review and gaps analysis. Int J Med Inform. Feb 2017;98:56–64. doi:10.1016/j.ijmedinf.2016.12.001

40. Jette DU, Hunter SJ, Burkett L, et al. Physical therapist management of total knee arthroplasty. Phys Ther. Aug 31 2020;100(9):1603–1631. doi:10.1093/ptj/pzaa099

41. Chen W, O’Bryan CM, Gorham G, et al. Barriers and enablers to implementing and using clinical decision support systems for chronic diseases: A qualitative systematic review and meta-aggregation. Implement Sci Commun. Jul 28 2022;3(1):81. doi:10.1186/s43058-022-00326-x

42. Van de Velde S, Heselmans A, Delvaux N, et al. A systematic review of trials evaluating success factors of interventions with computerised clinical decision support. Implement Sci. Aug 20 2018;13(1):114. doi:10.1186/s13012-018-0790-1

43. Horsky J, Schiff GD, Johnston D, Mercincavage L, Bell D, Middleton B. Interface design principles for usable decision support: A targeted review of best practices for clinical prescribing interventions. J Biomed Inform. Dec 2012;45(6):1202–16. doi:10.1016/j.jbi.2012.09.002

44. Bates DW, Kuperman GJ, Wang S, et al. Ten commandments for effective clinical decision support: Making the practice of evidence-based medicine a reality. J Am Med Inform Assoc. Nov-Dec 2003;10(6):523–30. doi:10.1197/jamia.M1370

45. Sutton RT, Pincock D, Baumgart DC, Sadowski DC, Fedorak RN, Kroeker KI. An overview of clinical decision support systems: Benefits, risks, and strategies for success. NPJ Digit Med. 2020;3:17. doi:10.1038/s41746-020-0221-y

