## Supplementary Table 1 for "Implementation of a “people-like-me” tool for personalized rehabilitation after total knee arthroplasty: A mixed methods pilot study"

**Supplementary Table 1.** Fidelity checklist provided to clinicians for reference and used by research assistant for fidelity assessments.

| **Required Tasks** | **Complete** |
| --- | --- |
| ***During the patient’s visit:*** | |
| 1. Patient completes survey |  |
| 2. PT measures the patient’s knee flexion ROM |  |
| 3. PT measures the patient’s knee extension ROM |  |
| 4. PT measures the patient’s TUG |  |
| 5. Knee flexion measurement is input into the tool |  |
| 6. Knee extension measurement is input into the tool |  |
| 7. TUG measurement is input into the tool |  |
| 8. WOMAC pain score is input into the tool |  |
| 9. Graph PDFs are printed from the tool and obtained by the PT |  |
| 10. PT discusses the PDF printouts with the patient including how the patient is recovering relative to “people-like-them” |  |
| 11. PT discusses how the tool’s output relates to the patient’s plan of care (e.g., frequency/duration of care, treatment emphasis, rehab goals, etc.) |  |
| 12. PT answers patient’s questions about the PDF printouts and/or asks the patient if they have questions about the printout |  |
| 13. PDF printout is sent home with the patient |  |
| **Total Complete** | /13 |
