## Supplementary Table 2 for "Implementation of a “people-like-me” tool for personalized rehabilitation after total knee arthroplasty: A mixed methods pilot study"

**Supplementary Table 2.** Description of PLM tool implementation strategy using Expert Recommendations for Implementing Change (ERIC) definitions

| **ERIC Strategy** | **ERIC Definition** | **How strategy was applied** |
| --- | --- | --- |
| Audit and provide feedback | Collect and summarize clinical performance data over a specified time period and give it to clinicians and administrators to monitor, evaluate, and modify provider behavior. | The participating clinics shared data related to implementation with the research team. The research team analyzed the data and shared the results with clinic leadership to identify potential sources of implementation challenges. |
| Develop and implement tools for quality monitoring | Develop, test, and introduce into quality-monitoring systems the right input—the appropriate language, protocols, algorithms, standards, and measures (of processes, patient/consumer outcomes, and implementation outcomes) that are often specific to the innovation being implemented. | The research team developed a list of core components for using the PLM tool. This list was shared with clinicians, and the research assistant conducted periodic fidelity assessments using the list. |
| Provide local technical assistance | Develop and use a system to deliver technical assistance focused on implementation issues using local personnel. | The local research assistant served as clinicians' point of contact for any technical or logistical issues related to using the PLM tool. |
| Promote adaptability | Identify the ways a clinical innovation can be tailored to meet local needs and clarify which elements of the innovation must be maintained to preserve fidelity. | The clinics were provided with the core components for implementing the PLM tool, but they were encouraged to adapt their own strategies for integrating these components within their workflow. |
| Build a coalition | Recruit and cultivate relationships with partners in the implementation effort. | The research team has collaborated with the participating clinics for nearly a decade in quality improvement and research efforts related to joint replacement. |
| Conduct educational meetings | Hold meetings targeted toward different stakeholder groups (e.g., providers, administrators, other organizational stakeholders; and community, patient/consumer, and family stakeholders) to teach them about the clinical innovation. | The research held educational meetings with each clinic before the pilot started and yearly thereafter. The meetings focused on (a) reviewing the project’s objectives, (b) reviewing the clinics’ planned implementation strategy, (c) troubleshooting any implementation problems, and (d) cultivating relationships with clinical staff |
| Develop educational materials | Develop and format manuals, toolkits, and other supporting materials in ways that make it easier for stakeholders to learn about the innovation and for clinicians to learn how to deliver the clinical innovation. | The research team developed online, self-paced educational materials that focused on (1) the tool’s purpose and how to use its web-based interface, (2) how to interpret “people-like-me” reference charts and how to engage patients with them, and (3) how to use information from the tool to inform personalized clinical decision making. The training also included quizzes to assess and reinforce clinician knowledge |
| Distribute educational materials | Distribute educational materials (including guidelines, manuals and toolkits) in person, by mail, and/or electronically. | Education materials were available on-demand through the clinics' established online training platform |
