## Supplementary File 1 for "Implementation of a “people-like-me” tool for personalized rehabilitation after total knee arthroplasty: A mixed methods pilot study"

**Supplementary File 1.** Adapted surveys to assess clinicians’ perceptions of PLM tool

1. **Acceptability of Intervention Measure^1^**

We are interested to know more about your experience using the PLM tool with patients after total knee replacement.

**Please circle only one answer for each statement.** There is no right or wrong answer for these questions. We are interested in your opinion based on your experience using the PLM tool.

| **Item** | **Completely disagree** | **Disagree** | **Neither agree nor disagree** | **Agree** | **Completely agree** |
| --- | --- | --- | --- | --- | --- |
| 1. The PLM tool meets my approval | 1 | 2 | 3 | 4 | 5 |
| 2. The PLM tool is appealing to me | 1 | 2 | 3 | 4 | 5 |
| 3. I like the PLM tool | 1 | 2 | 3 | 4 | 5 |
| 4. I welcome the PLM tool | 1 | 2 | 3 | 4 | 5 |

2. **Feasibility of Intervention Measure^1^**

We are interested to know more about your experience using the PLM tool with patients after total knee replacement.

**Please circle only one answer for each statement.** There is no right or wrong answer for these questions. We are interested in your opinion based on your experience using the PLM tool.

| **Item** | **Completely disagree** | **Disagree** | **Neither agree nor disagree** | **Agree** | **Completely agree** |
| --- | --- | --- | --- | --- | --- |
| 1. The PLM tool seems implementable | 1 | 2 | 3 | 4 | 5 |
| 2. Using the PLM tool seems possible | 1 | 2 | 3 | 4 | 5 |
| 3. Using the PLM tool seems doable | 1 | 2 | 3 | 4 | 5 |
| 4. The PLM tool seems easy to use | 1 | 2 | 3 | 4 | 5 |

3. **System Usability Scale^2^**

We are interested to know more about your experience using the PLM tool with patients after total knee replacement.

**Please circle only one answer for each statement.** There is no right or wrong answer for these questions. We are interested in your opinion regarding the PLM tool’s usability.

|  | **Item** | **Strongly disagree** |  |  |  | **Strongly agree** |
| --- | --- | --- | --- | --- | --- | --- |
|  | 1. I think that I would like to use the PLM tool frequently | 1 | 2 | 3 | 4 | 5 |
|  | 2. I found the PLM tool to be unnecessarily complex | 1 | 2 | 3 | 4 | 5 |
|  | 3. I thought the PLM tool was easy to use | 1 | 2 | 3 | 4 | 5 |

**Supplementary File 1 continued.**

| **Item** | **Strongly disagree** |  |  |  | **Strongly agree** |
| --- | --- | --- | --- | --- | --- |
| 4. I think that I would need the support of a technical person to be able to use the PLM tool | 1 | 2 | 3 | 4 | 5 |
| 5. I found the various functions in the PLM tool were well integrated | 1 | 2 | 3 | 4 | 5 |
| 6. thought there was too much inconsistency in the PLM tool | 1 | 2 | 3 | 4 | 5 |
| 7. I would imagine that most people would learn to use the PLM tool very quickly | 1 | 2 | 3 | 4 | 5 |
| 8. I found the PLM tool to be very cumbersome to use | 1 | 2 | 3 | 4 | 5 |
| 9. I felt very confident using the PLM tool | 1 | 2 | 3 | 4 | 5 |
| 10. I needed to learn a lot of things before I could get going with the PLM tool | 1 | 2 | 3 | 4 | 5 |

4. **Perceived Characteristics of Intervention Scale^3^**

We are interested to know more about your perception of the PLM tool. There are no right or wrong answers for these questions.

| **Item** | **Strongly disagree 1** | **Disagree 2** | **Neither agree nor disagree 3** | **Agree 4** | **Strongly Agree 5** |
| --- | --- | --- | --- | --- | --- |
| 1. The PLM tool is more effective than other methods I have used to monitor recovery after knee replacement |  |  |  |  |  |
| 2. The PLM tool is more convenient than other methods I have used to monitor patient recovery after knee replacement |  |  |  |  |  |
| 3. Using the PLM tool fits well with the way I like to work |  |  |  |  |  |
| 4. The PLM tool is aligned with my clinical judgment |  |  |  |  |  |
| 5. The PLM tool is clear and understandable |  |  |  |  |  |
| 6. The PLM tool is easy to use |  |  |  |  |  |

**Supplementary File 1 continued.**

| **Item** | **Strongly disagree 1** | **Disagree 2** | **Neither agree nor disagree 3** | **Agree 4** | **Strongly Agree 5** |
| --- | --- | --- | --- | --- | --- |
| 7. The PLM tool can be tested out with patient without disrupting their overall therapy |  |  |  |  |  |
| 8. It is easy to try out the PLM tool with patients without disrupting their overall therapy |  |  |  |  |  |
| 9. It is easy to tell whether patients are benefitting from the PLM tool |  |  |  |  |  |
| 10. The PLM tool produces improvements in my patients that I can actually see |  |  |  |  |  |
| 11. The PLM tool can be adapted to fit my treatment setting |  |  |  |  |  |
| 12. The PLM tool can be adapted to meet the needs of my patients |  |  |  |  |  |
| 13. Using the PLM tool improves the quality of the work that I do |  |  |  |  |  |
| 14. Using the PLM tool makes it easier to do my job |  |  |  |  |  |
| 15. The knowledge required to learn the PLM tool can be effectively taught |  |  |  |  |  |
| 16. The skills required to learn the PLM tool can be effectively taught |  |  |  |  |  |
| 17. The PLM tool training materials are helpful |  |  |  |  |  |
| 18. The PLM tool has helpful supportive materials for patients |  |  |  |  |  |

1. 1. Weiner BJ, Lewis CC, Stanick C, et al. Psychometric assessment of three newly developed implementation outcome measures. *Implement Sci*. Aug 29 2017;12(1):108. doi:10.1186/s13012-017-0635-3
2. 2. Brooke J. Sus-a quick and dirty usability scale. *Usability evaluation in industry*. 1996;189(194):4-7.
3. 3. Cook JM, Thompson R, Schnurr PP. Perceived characteristics of intervention scale: Development and psychometric properties. *Assessment*. Dec 2015;22(6):704-14. doi:10.1177/1073191114561254
